# COVID-19 dynamics across the US: A deep learning study of human mobility and social behavior

**DOI:** 10.1101/2020.09.20.20198432

**Authors:** Mohamed Aziz Bhouri, Francisco Sahli Costabal, Hanwen Wang, Kevin Linka, Mathias Peirlinck, Ellen Kuhl, Paris Perdikaris

## Abstract

This paper presents a deep learning framework for epidemiology system identification from noisy and sparse observations with quantified uncertainty. The proposed approach employs an ensemble of deep neural networks to infer the time-dependent reproduction number of an infectious disease by formulating a tensor-based multi-step loss function that allows us to efficiently calibrate the model on multiple observed trajectories. The method is applied to a mobility and social behavior-based SEIR model of COVID-19 spread. The model is trained on Google and Unacast mobility data spanning a period of 66 days, and is able to yield accurate future forecasts of COVID-19 spread in 203 US counties within a time-window of 15 days. Strikingly, a sensitivity analysis that assesses the importance of different mobility and social behavior parameters reveals that attendance of close places, including workplaces, residential, and retail and recreational locations, has the largest impact on the basic reproduction number. The model enables us to rapidly probe and quantify the effects of government interventions, such as lock-down and re-opening strategies. Taken together, the proposed framework provides a robust workflow for data-driven epidemiology model discovery under uncertainty and produces probabilistic forecasts for the evolution of a pandemic that can judiciously inform policy and decision making. All codes and data accompanying this manuscript are available at https://github.com/PredictiveIntelligenceLab/DeepCOVID19.

## 1 Introduction

Since its emergence in late December 2020 [1], as of early September 2020, the novel SARS-CoV-2 virus (COVID-19) has resulted in over 30 million of confirmed cases globally, of which nearly 1 million of people have died [2]. After several months, active cases still surge in many countries and regions [2]. To date, there are no known preventive pharmaceutical interventions [3], and numerous behavioral intervention policies, such as lock-down, mandatory self-isolation and facial covering orders [4] have been issued to reduce the transmission of such a highly infectious disease to an extent that current health and financial systems could sustain. Without these timely countermeasures, breakdown of these systems will lead to more severe social and economic crisis in an unexpectedly fast pace, especially given the exponential rise in cases number at the beginning of the outbreak as it was the case in the US, for instance for the first few months of the COVID-19 spread [5].

Despite their effectiveness in containing the aggressive virus spread, these attempts have also interrupted ordinary living orders, and have resulted in heavy socio-economical impacts such as exploding unemployment rates, prevailing emotional distress, and the consequently exacerbated civil unrest [6]. The phased re-opening strategy [7] adopted by many states and cities as a compromise, seemingly becomes the only way to leverage the casualties brought by the disease itself, and by the consequential economic interruption. Therefore, accurately and quantitatively assessing the cost and benefit of restricted opening gains an increasing importance nowadays, especially at a time when America saw its second spike in tested positive population after some controversial policies.

Studies [8, 9] have revealed that the transmission of COVID-19 likely comes from direct contact, such as droplets from infected persons, or indirect contact, where virus that remained active on shared surface is brought into human by body means of involuntary behaviors such as whipping face with hands [10]. After entering the airway system, the virus replicates itself in the throat, and further causes lung lesions. The incubation period before the infection triggers human immune responses, most commonly fever and dry cough, could last 6 days on average or even as long as 14 days observed [11]. During or even after the incubation period, the patients can still remain largely asymptomatic, while still being able to infect others [12] [13] [14]. The corresponding antigen detection assay using throat wash and saliva is also reported to produce false positive or negative result [15], affecting the reliability of the case data resource.

Such pathology makes clear that close contact interactions in public space is a major cause of the spread of COVID-19. Individual level transmission models [16] have shown that 80% of secondary transmissions may have been caused by a small fraction of infectious individuals (*∼*10%) from some super-spreading events, mainly large and dense gatherings. The aforementioned two challenges, long incubation period and noisy data, call for timely adjustments to existing policy plans based on robust assessments of the situation. Under such demand, cell phones, devices that most people carry almost all the time with capability of accurate positioning and behavioral recording, become an ideal carrier of information capturing the daily human migration patterns that could be used to estimate the magnitude of multiple dimensions of social mixing. For example, many educational institutions whose operations require gathering of a large number of people will begin mandating the use of mobile applications [17] for contact tracing purposes. Technology companies such as Google [18] and Apple [19] also make publicly available their privacy preserved databases containing mobility trend data ranging from working to recreational. These disaster countermeasures provide a firm ground of valuable information [**?**], which this study sets to explore in order to investigate the predictability of COVID-19 dynamics, and its dependence on human mobility and social behavior patterns.

Specifically, here we conduct a large-scale computational study that leverages Google’s mobility trend data [18] around grocery and pharmacy stores, parks, public transportation, retail and recreation spaces, residential and workplaces, along with the Unacast’s social behavior data [20] which includes information on the average distance traveled, visits to non-essential retail and services, and unique human encounters per km^2^ relative to national metrics pre-COVID-19. We believe that these types of mobility and social behavior trends cover most scenarios of social mixing and can be used as predictive indicators of viral spread [21]. The dynamic recordings are normalized by the baseline mobility trend value, calculated with data from January, 2020.

Similar to other big-data type data sources, the mobility and social behavior trend data suffer from a fair amount of noise, which becomes one of the challenges that we wish to address in our framework. Another key challenge associated with the accurate calibration of disease spread models is the availability of reliable data. One of the simplest compartmental models in epidemiology, the SIR model [22], includes three groups of individuals among a population: susceptible, infected, and recovered or deceased individuals. However, reliable data is only available for the cumulative number of the infected population. This difficulty is further pronounced for more complicated models accounting for the individuals who have been infected but are not yet infectious themselves (SEIR model), for asymptomatic cases [23], [24] or for the effect of government policies [25]. From a mathematical viewpoint, this requires us to calibrate a large number of parameters that characterize a complex nonlinear dynamical system using partial and incomplete observations. Moreover, due to the inherent uncertainty related to the data and to compartment models themselves, a probabilistic characterization of the estimated parameters is required such that extrapolations in time can be provided with quantified uncertainty.

From a modeling perspective, the main novelty of this work stems from utilizing mobility and social behavior trend data to model the temporal evolution of the COVID-19 infectious rate - the probability of transmitting disease between a susceptible and an infectious individual (Figure 1). From a methodological standpoint, this work generalizes the multi-step neural network method of Raissi *et al*. [26] to enable gradient-based optimization of unknown epidemiology model parameters with latent variables. This is achieved by effectively back-propagating the influence of the unobserved variables through the disease spread dynamics with an appropriate tensorization of the loss function. This key step enables a computationally efficient treatment of multiple temporal trajectories, as well as the concurrent training of multiple neural networks. The latter, allows us to generate a statistical ensemble over the predicted evolution of the underlying disease spread dynamics, which enables future forecasting with quantified uncertainty. In summary, our specific contributions can be organized as follows:

**Figure 1:**
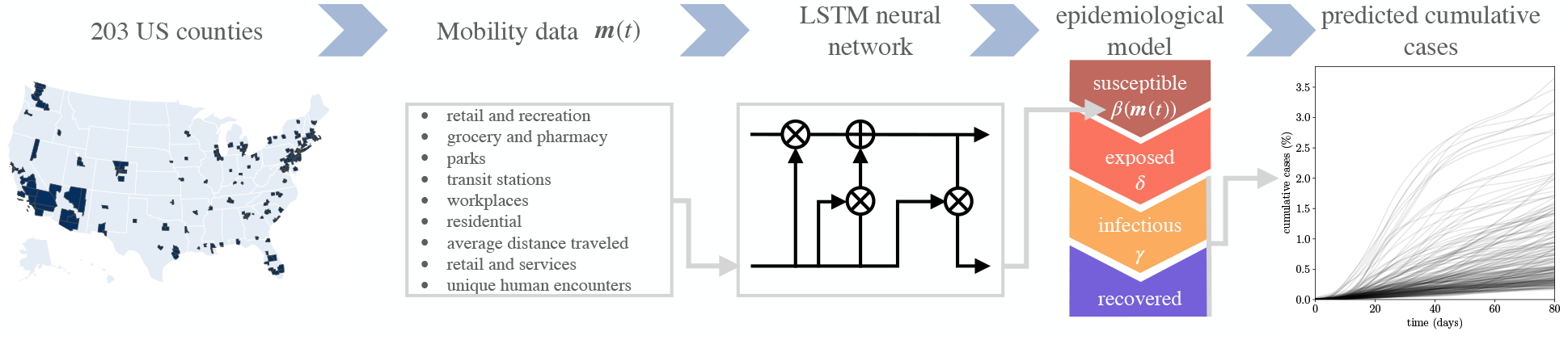
Our goal is to develop a robust computational framework for calibrating COVID-19 models from noisy time-series data. This will enable the quantitative assessment of government policies (e.g., travel bans, quarantine, contact tracing, etc.), and yield reliable forecasts with quantified uncertainty that can help us manage and contain an epidemic.

- We extend the multi-step neural network method of Raissi *et. al*. [26] to enable dynamical system’s parameters inference with unobserved latent variables. Moreover, we tensorize the loss function formulation in order to perform the parameters inference from multiple trajectories of different counties, while having a small memory footprint in terms of the size of the associated computational graph. Hence, the proposed approach is computationally efficient and enables training of multiple neural networks, which provides an uncertainty quantification estimate of the dynamical system evolution via multiple trajectory forecasts.
- We model the basic reproduction number with a long short-term memory neural network (LSTM) to capture its temporal dependence on the mobility and social behavior variables. The LSTM architecture enables to account for the time lag between the change in mobility and social behavior, and the effect on the spread of the disease.
- We demonstrate the effectiveness of the proposed method by applying it to the COVID-19 spread in 203 US counties such that the training is performed using data for a period of 66 days, and we produce reliable future forecasts with quantified uncertainty for a period of 15 days. The lock-down efficiency is quantified by the subsequent forecasted decrease of the basic reproduction number. We also perform a sensitivity analysis showing that among the mobility parameters, the attendance of closed places has the highest influence on the evolution of the basic reproduction number, unlike open or transit locations. Moreover, all social behavior parameters have relatively important sensitivities, highlighting the importance of social interactions on the spread of the virus.

Our findings put forth a novel, flexible and robust workflow for data-driven epidemiology model discovery that can potentially help health authorities take a more quantitative and judicious approach to policy making in order to slow down COVID-19 propagation. We demonstrate that well-calibrated epidemiology models can indeed return sensible future forecasts. Such capability can directly assist with optimizing resource allocation for more precise diagnostics and treatment, as well as yield a better understanding of the disease spread dynamics. Taken together, this can be another tool in our arsenal to help us better prepare for COVID-19 and potential future pandemics, and provide additional quantitative tools to help us minimize their spread.

This paper is organized as follows. Section 2.1 presents the mobility and social behavior-based SEIR epidemiology model considered to study the spread of COVID-19. Section 2.2 details the proposed computational method for inference of the mobility and social behavior-based SEIR model parameters and the corresponding technical ingredients. In section 3, the effectiveness of the proposed method is demonstrated by testing it on real mobility and infections data from 203 US counties and obtaining reliable future forecasts for a period of 15 days with quantified uncertainty. Within the same section, we present a sensitivity analysis to assess the influence of the different mobility and social behavior parameters considered in the spread of the disease. Finally, in section 4 we summarize our key findings, discuss the limitations of the proposed approach, and carve out directions for future investigation.

## 2 Methods

### 2.1 Epidemiology Model

We consider a mobility and social behavior-based SEIR model detailed as follows:

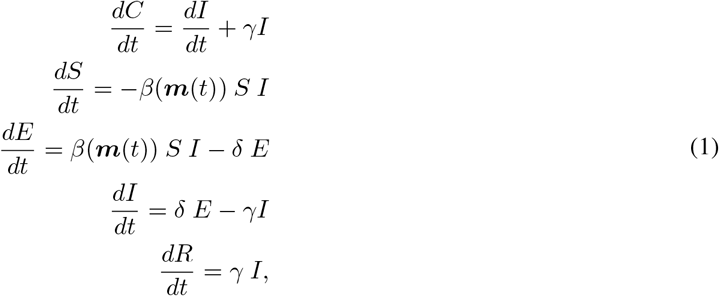

where *S* is the fraction of susceptible population, *E* the fraction of exposed population including individuals who have been infected but are not yet infectious themselves during the incubation period, *I* the fraction of infectious population, *R* the fraction of removed population that accounts for recovered and deceased individuals, and *C*(*t*) *≡R*(*t*) + *I*(*t*) is the fraction of cumulative cases, which is the only variable with available data over time. Note that we consider an SEIR model with variables normalized by the number of population such that the resulting model (1) is universal for all regions and independent of the number of their inhabitants.

Since the spread of an infectious disease that is transmitted from human to human depends on interpersonal interactions, we model the susceptible to infectious transition rate *β* in equation (1) as a function of mobility and social behavior parameters denoted as 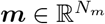, which contains the mobility and social behavior parameters 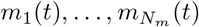. This enables determining the evolution of the basic reproduction number, 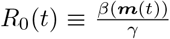, a key paremeter to characterize the evolution of an epidemic.

The proposed model is trained on data provided by Google for mobility change from baseline [18] (parameters *m*_1_, …, *m*_6_ as discussed below), along with the data provided by Unacast [20] for social behavior trends (parameters *m*_7_, …, *m*_9_ as detailed below) for all 203 US counties considered in this work. Specifically, we consider a total of *N*_*m*_ = 9 mobility and social behavior parameters corresponding to:

- *m*_1_: retail and recreation percent change from baseline,
- *m*_2_: grocery and pharmacy percent change from baseline,
- *m*_3_: parks percent change from baseline,
- *m*_4_: transit stations percent change from baseline,
- *m*_5_: workplaces percent change from baseline,
- *m*_6_: residential percent change from baseline,
- *m*_7_: average distance traveled,
- *m*_8_: visits to non-essential retail and services,
- *m*_9_: unique human encounters per Km^2^ relative to national pre-Covid-19.

Figure 2 provides the observed mobility and social behavior trends for three counties randomly selected among the data-set considered, showing the inherent presence of noise within the trajectories.

**Figure 2:**
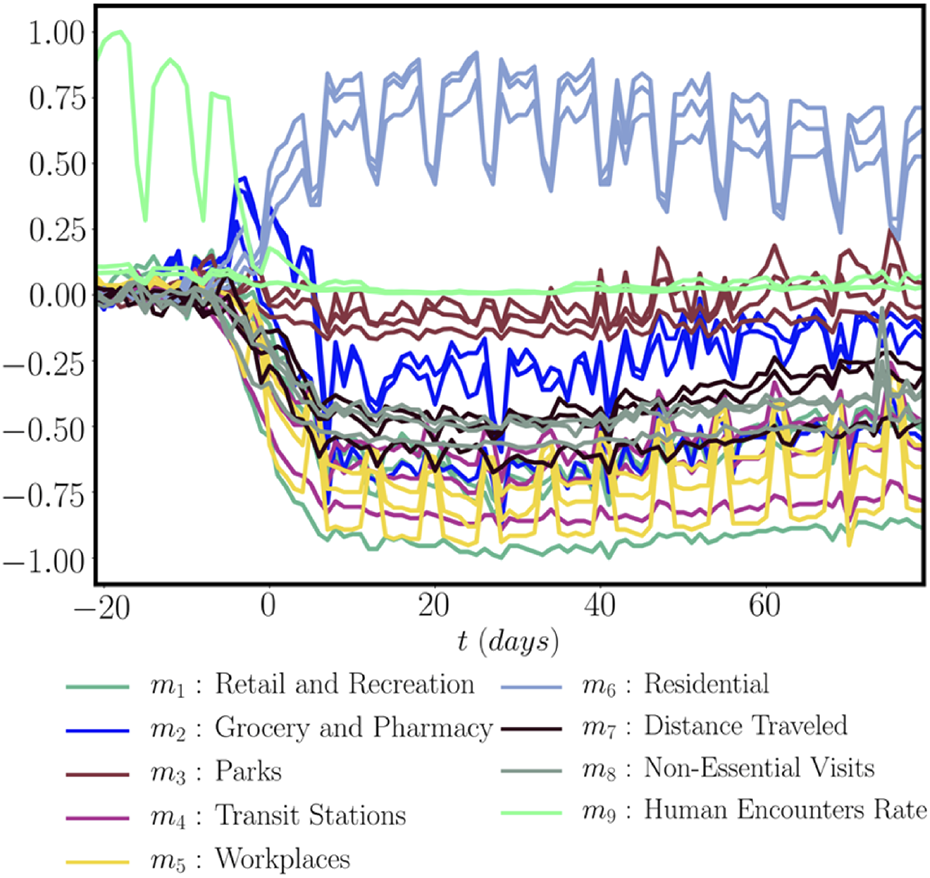
Mobility and social behavior parameters for three counties randomly sampled among the data-set considered.

### 2.2 A deep learning approach to model calibration given partially observed states

In this work, *γ* and *δ* are assumed as given and equal to 1*/*6.5 [27, 28] and 1*/*2.5 [27, 11] respectively for COVID-19, and the task is to learn the dynamics of *β*(*t*), and, consequently, the evolution of the solution to the SEIR model of equation (1). The learning task can then be formulated as follows.

Given *γ, δ* and a data-set 𝒟 for *N*_*c*_ different regions containing: (i) the initial conditions of the variable *I*(*t* = *t*_0_) for each region, (ii) the time trajectories of the observed variable *C*(*t*) for each region, and (iii) the time trajectories of the observed mobility and social behavior trends ***m***(*t*) for each region, infer the dependence of *β*(*t*) *≡ β*(***m***(*t*)) and the initial fractions of the exposed population *E*(*t* = *t*_0_) for all the different regions.

The observed data-set *𝒟* can be expressed as follows:

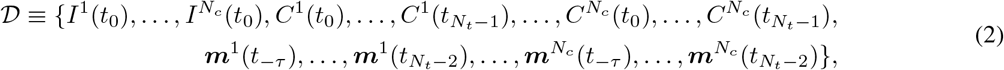

where superscripts refer to the different regions considered. In order to learn the mapping *β ≡ β*(***m***(*t*)), *β* is modeled with a long-short term memory (LSTM) [29] neural network of the form

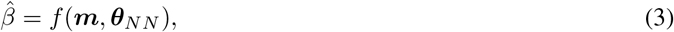

where ***θ***_*NN*_ refers to the weights and bias parameters defining the LSTM network. Since we are modeling the spread of an infectious disease with an incubation period, we utilize the lag structure of the LSTM network to model *β* as a function of the mobility and social behavior parameters over a period of time of *τ* days. The forward pass of LSTM neural network considered in this work is detailed in appendix A.

Given the available dataset 𝒟, we formulate a multi-step loss function in order to infer the learnable parameters ***θ*** given in equation (4), which include both the LSTM parameters ***θ***_*NN*_ as well as the latent initial fractions of the exposed population for all *N*_*c*_ different regions considered,

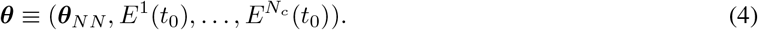

To formulate the multi-step loss function, we follow the approach put forth by Raissi *et al*. [26], and consider an appropriate temporal discretization of equation (1. Given the available data containing the initial conditions of the variable *I*(*t* = *t*_0_) and the trajectories of the variable *C*(*t*), the variable *I*(*t*) can be estimated for the different time instances 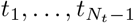 using the first equation of the SEIR model (1). For instance, using the trapezoidal discretization rule [26, 30], the first equation of the SEIR model (1) for region *j* gives

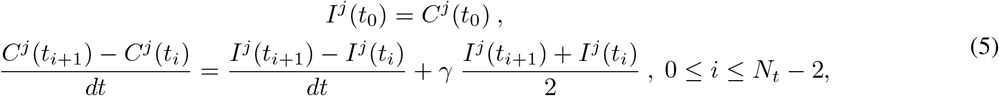

where *dt* is the time-step size corresponding to the period of the infections number update, and taken equal to 1 day in this work based on the available data. Then, given the estimated *I*(*t*) from equation (5) and the parameters ***θ***, we can compute *Ŝ*(*t*), then *Ê*(*t*) and finally *Î*(*t*) using the remaining equations of the SEIR model (1). Using the trapezoidal rule for instance, such discretization for region *j* gives:

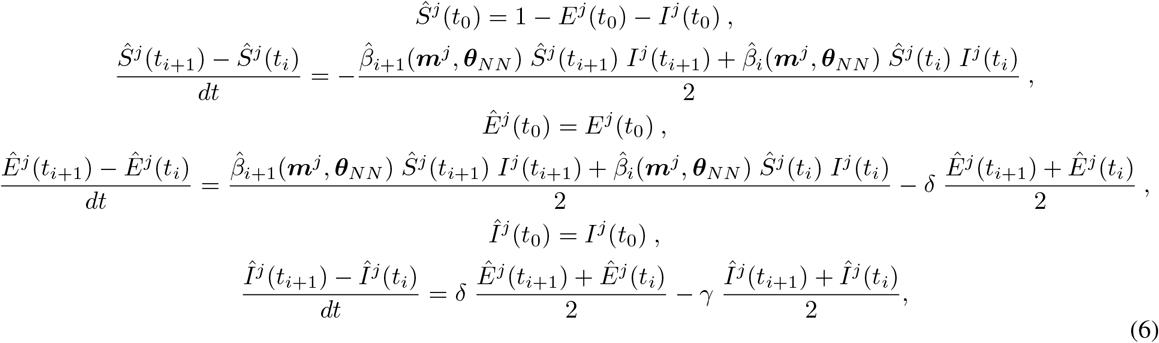

where 0 ≤ *i* ≤ *N*_*t*_ − 2 and 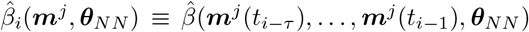 is the LSTM prediction of *β* for region *j* at time instance *t*_*i*_. Note that *E*^*j*^(*t*_0_) appearing in the initial conditions in equations (6) is part of the learnable parameters ***θ***, as described in equation (4). Notice that, given the dataset 𝒟 and the parameters ***θ***, one can estimate *Ŝ*(*t*), *Ê*(*t*), and *Î*(*t*) in that order. Finally, the loss function for a single neural network is expressed as the difference between the variable *I*(*t*) estimated only from the available data and the estimated variable *Î*(*t*) which depends on the learnable parameters

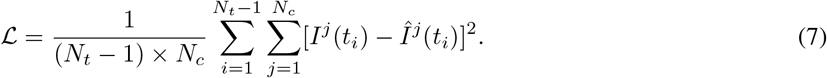

One crucial point in terms of implementation consists of the possibility of expressing the finite difference equation of (5), and each of the finite difference equations of (6), as tensors of shape *N*_*c*_ × (*N*_*t*_ − 1). As a consequence, the loss given in equation (7) can be expressed as the sum of the elements of a tensor of shape *N*_*c*_ × (*N*_*t*_ − 1) ensuring a minimalist computational graph, which speeds up gradient back-propagation. We emphasize that the proposed methodology is independent of the chosen finite difference scheme, and the trapezoidal rule is chosen here for sake of presentation and in coherence with the finite difference scheme adopted in this work as justified in appendix A. This computational strategy allows us to consider multiple trajectories simultaneously (accounting for the different regions considered in the data), instead of performing the learning task using observations from a single or a handful of trajectories. Finally, the loss can be further tensorized to enable the parallel training of multiple neural networks. By training an ensemble of independent networks, we can obtain a statistical characterization of model uncertainty and quantify its effect on the resulting model predictions and forecasts. For more technical details, the interested reader is referred to our publicly available implementation^1^.

## 3 Results

All code and data presented in this section will be made publicly available at https://github.com/ PredictiveIntelligenceLab/DeepCOVID19. The minimization of equation (7) is carried out using stochastic Adam updates [31] with the hyper-parameters summarized in table 1. Based on our experience, the reported results are robust with respect to the numerical values chosen for the different optimization hyper-parameters.

**Table 1:** Hyper-parameter settings employed across all numerical studies.

|  |  |
| --- | --- |
| $\theta_{NN}$ initialization | Xavier initialization |
| $E^1(t_0), \dots, E^{N_c}(t_0)$ initialization | Uniform initialization |
| Optimization algorithm | Adam |
| Learning rate | $10^{-3}$ |
| Number of iterations | $2 \times 10^4$ |
| County batch size | 60 |

### 3.1 Training data and pre-processing

We remove the counties containing missing mobility or social behavior parameters for more than 3 consecutive days or 20 days in total over the considered time period, and we use linear interpolation for missing data for 3 or less consecutive days. We also consider the USAFacts processed infections data for all US counties [32]. The two datasets are cross-matched together such that the considered counties satisfy the following criteria:

- the infection rate *C*(*t*) is at least equal to 0.2% after *t* = 81 days of the spread of the disease,
- the mobility and social behavior data for the *τ* days prior to the first day of infections is available.

For each county, the first day of infections is defined as the first day for which there are at least 10 infections. These criteria provides a dataset with 203 US counties and a period of time of 81 days, such that the first day of infections for each county is between March 16, 2020 and March 24, 2020, while the last day of considered is between June 4, 2020 and June 12, 2020. The total population of these counties is 150 million, which represents 46% of total US population.

### 3.2 Sample Trajectories with Forecasts

The proposed framework is trained on the collected data corresponding to the first 66 days. For the remaining 15 days, only the mobility and social behavior data are used in order to extrapolate the fraction of infected population. We train an ensemble of 100 LSTM networks with randomized parameter initializations. The total training time takes 14.7 minutes on a single NVIDIA Tesla P100 GPU. Once trained, the model provides estimates of the cumulative cases fraction with a relative error equal to 4.3% on the training data (for the first 66 days), and a relative error equal to 6.6% on the testing data, corresponding to the extrapolation carried out for the remaining 15 days of the data-set. Figure 3 shows the distribution of relative errors averaged over the 15 days extrapolation window across all US counties considered. Evidently, the proposed model can yield accurate future forecasts of COVID-19 spread, particularly for counties having a higher fraction of cumulative cases. However, using an SEIR model with a constant *β* over time for each county fails to explain the spread of COVID-19 for the 203 US counties considered. Figure 13 in appendix B compares the relative forecasting errors averaged over the extrapolation period when using the proposed multi-step LSTM approach and a multi-step approach with a constant reproduction number. Based on these results we conclude that enhancing the expressiveness of the SEIR model via the inclusion of mobility and social behavior-based effects in the time-depended modeling of *β* has a significant impact on the generalization accuracy of the model, leading to more accurate future forecasts.

**Figure 3:**
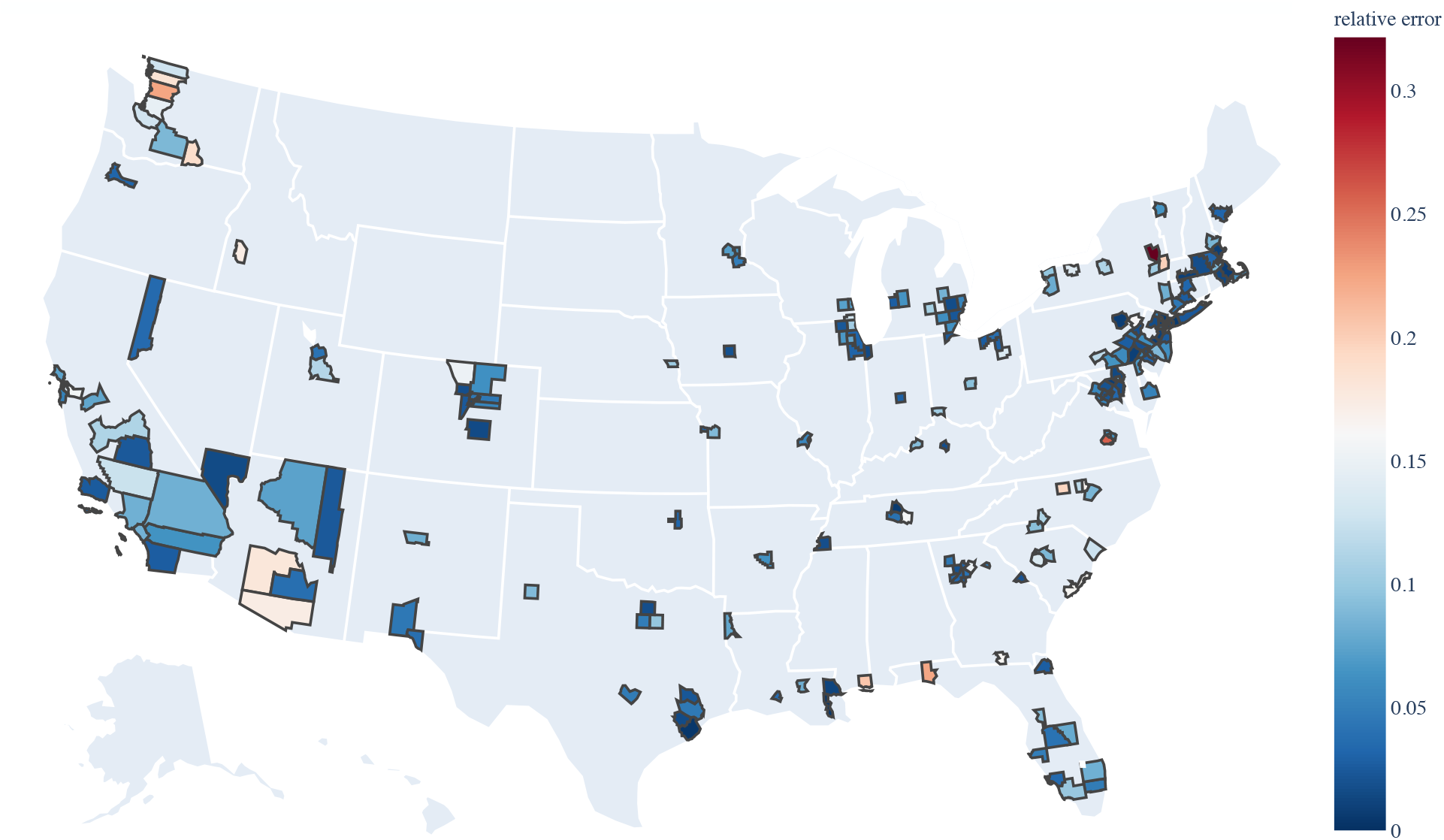
Relative forecasting errors averaged over the 15 days extrapolation period in all 203 US counties considered in this work.

**Figure 13:**
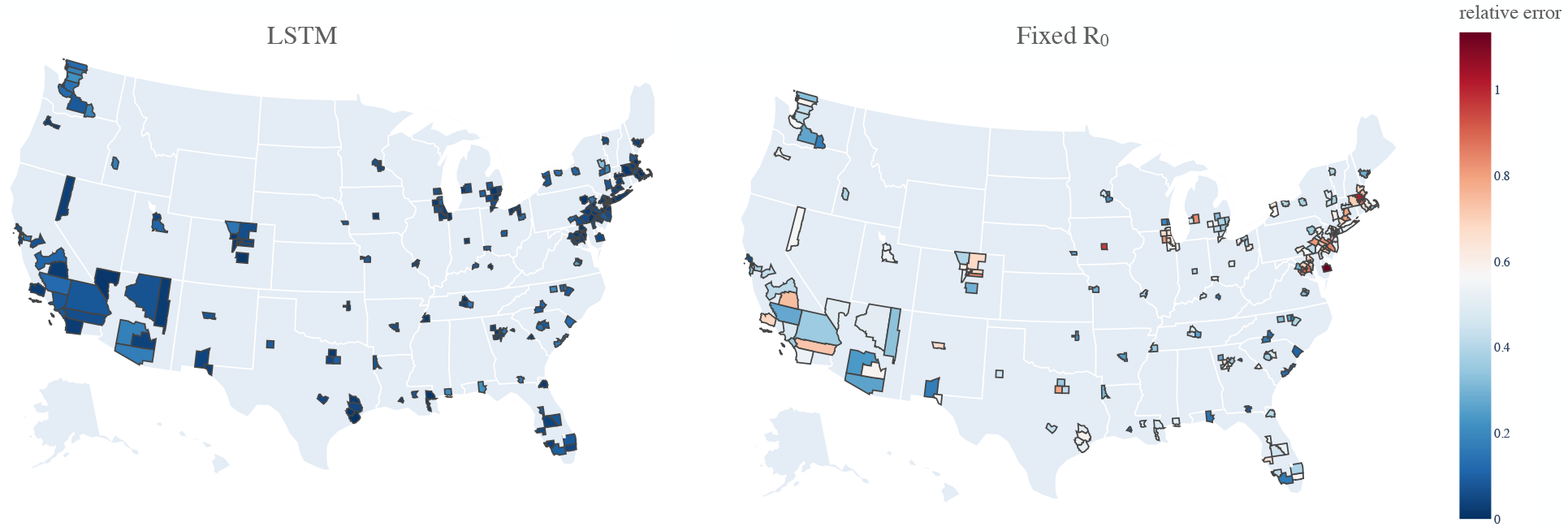
*Predictive error comparison:* Relative forecasting errors averaged over the 15 days extrapolation period in all 203 US counties considered in this work. (Left) Proposed multi-step LSTM approach modeling a time-dependent reproduction number *β*(***m***(*t*)), (right) a multi-step approach with a constant reproduction number *β*.

Figure 4 provides the dependency of the relative error averaged over the 15 days extrapolation on the fraction of cumulative cases on the last day. The latter shows a better accuracy for counties with higher cumulative cases fractions, which can be explained by the loss function considered (7). Indeed, counties with higher fraction of cumulative cases present a higher fraction of the infectious population. As a consequence, the corresponding terms within the loss function are generally higher than the ones related to counties with a lower fraction of cumulative cases. Hence the learning of the model is inherently more driven towards minimizing the errors of the estimates related to counties with higher fraction of cumulative cases, which explains the lower errors observed compared to counties with lower fraction of cumulative cases. One possibility to remedy to this issue is to consider normalized errors in the loss function (7) by dividing the L2 errors [*I*^*j*^(*t*_*i*_)−*Î*^*j*^ (*t*_*i*_)]^2^ by the values of *I*^*j*^(*t*_*i*_), 1 ≤ *i* ≤ *N*_*t*_−1, 1 ≤ *j* ≤ *N*_*c*_. However, such approach would drive the model to better approximate counties with lower fraction of cumulative cases due to the importance weight introduced by the corresponding small denominator *I*^*j*^(*t*_*i*_). Since accurately modeling counties with higher disease propagation, and hence higher fraction of cumulative cases, is more crucial than focusing on counties with lower infection rates, we opted for the non-normalized loss function. Note that considering the normalized loss and adding a hyper-parameter within the denominator may partially solve the issue and slightly remedy the disparity in accuracy, however the improvement observed was not significant and it would make the proposed approach a problem-dependent method since the optimal hyper-parameter introduced would differ depending on the data-set considered.

**Figure 4:**
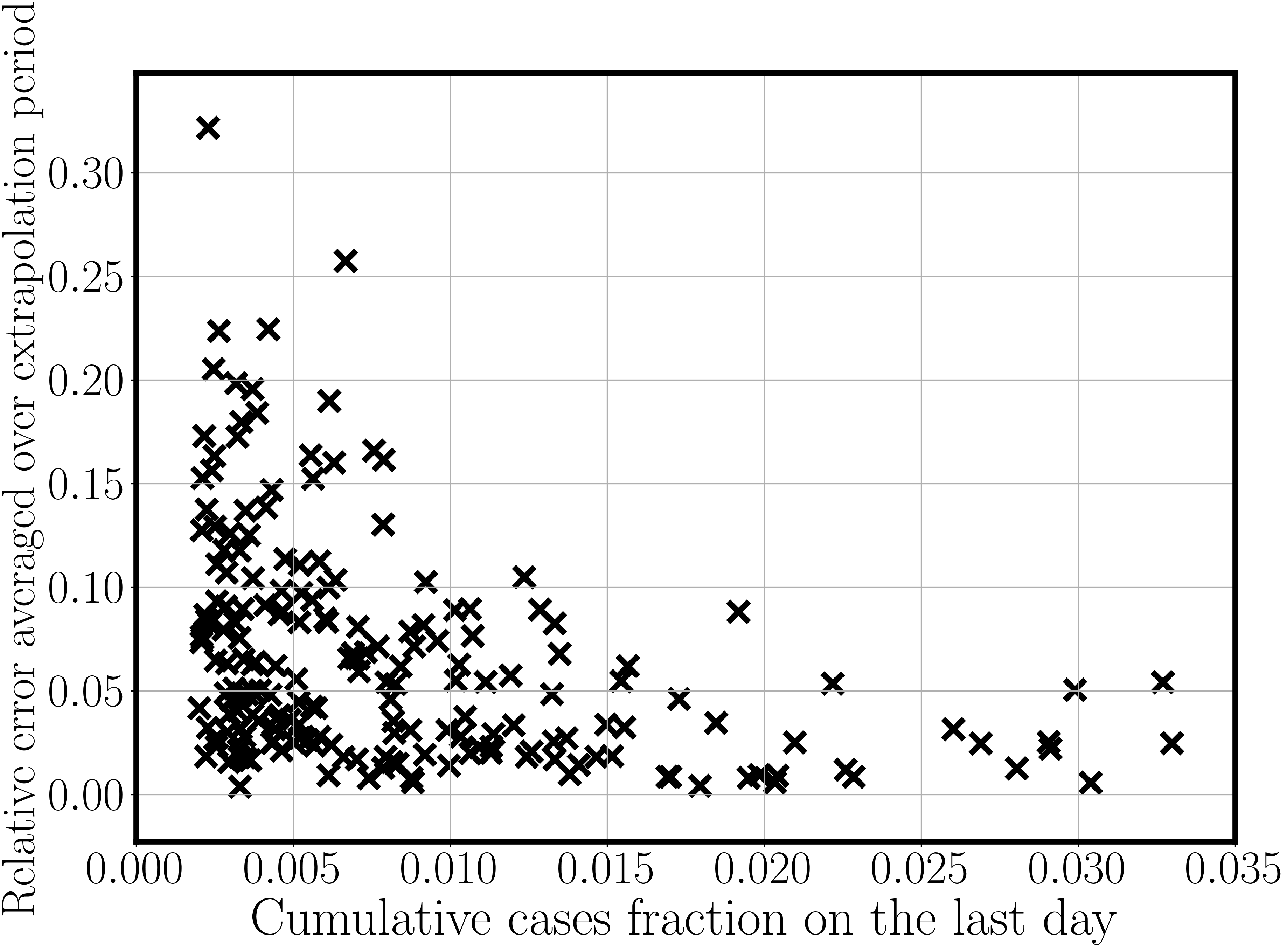
Variation of model accuracy as a function of the cumulative cases fraction.

New York state has the highest number of counties with a fraction of infected population above 1% after 81 days of the spread of the virus. Figure 5 shows the predictions obtained for the evolution of the fraction of infected population for the major New York counties. Figures 6 and 7 provide additional sample results for other US counties. New Jersey and Massachusetts present the second and third highest number of counties with a fraction of infected population above 1% after 81 days of the spread of the virus.

**Figure 5:**
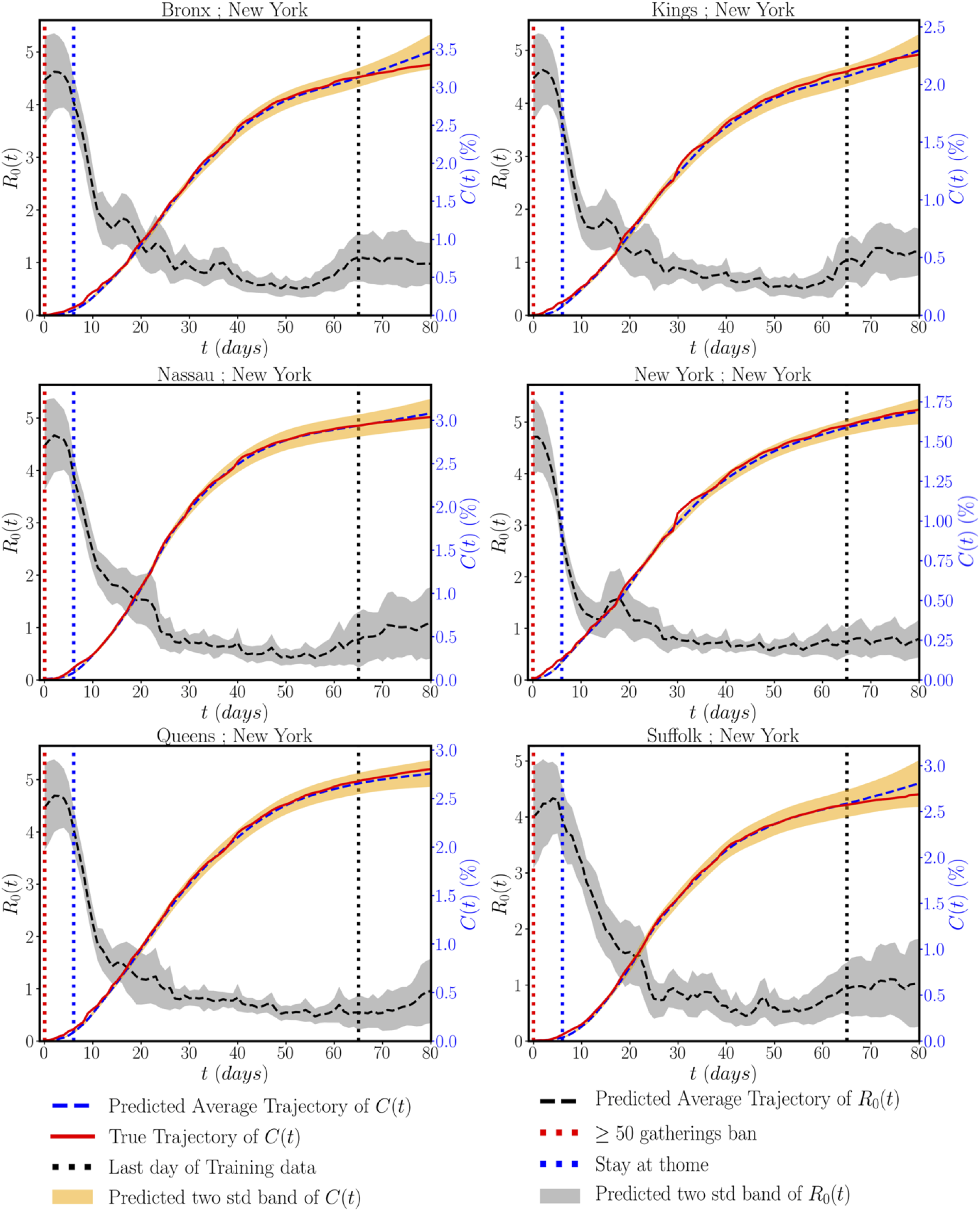
Evolution of the fraction of cumulative cases *C*(*t*) and the basic reproduction number *R*_0_(*t*) across different New York state counties. The red and blue horizontal dashed lines indicate the dates corresponding to the large crowd gathering ban and stay at home order issued by each county. The black horizontal dashed line delineates the data used for training the model (days 0-65), and the predicted model forecasts (days 66-80).

**Figure 6:**
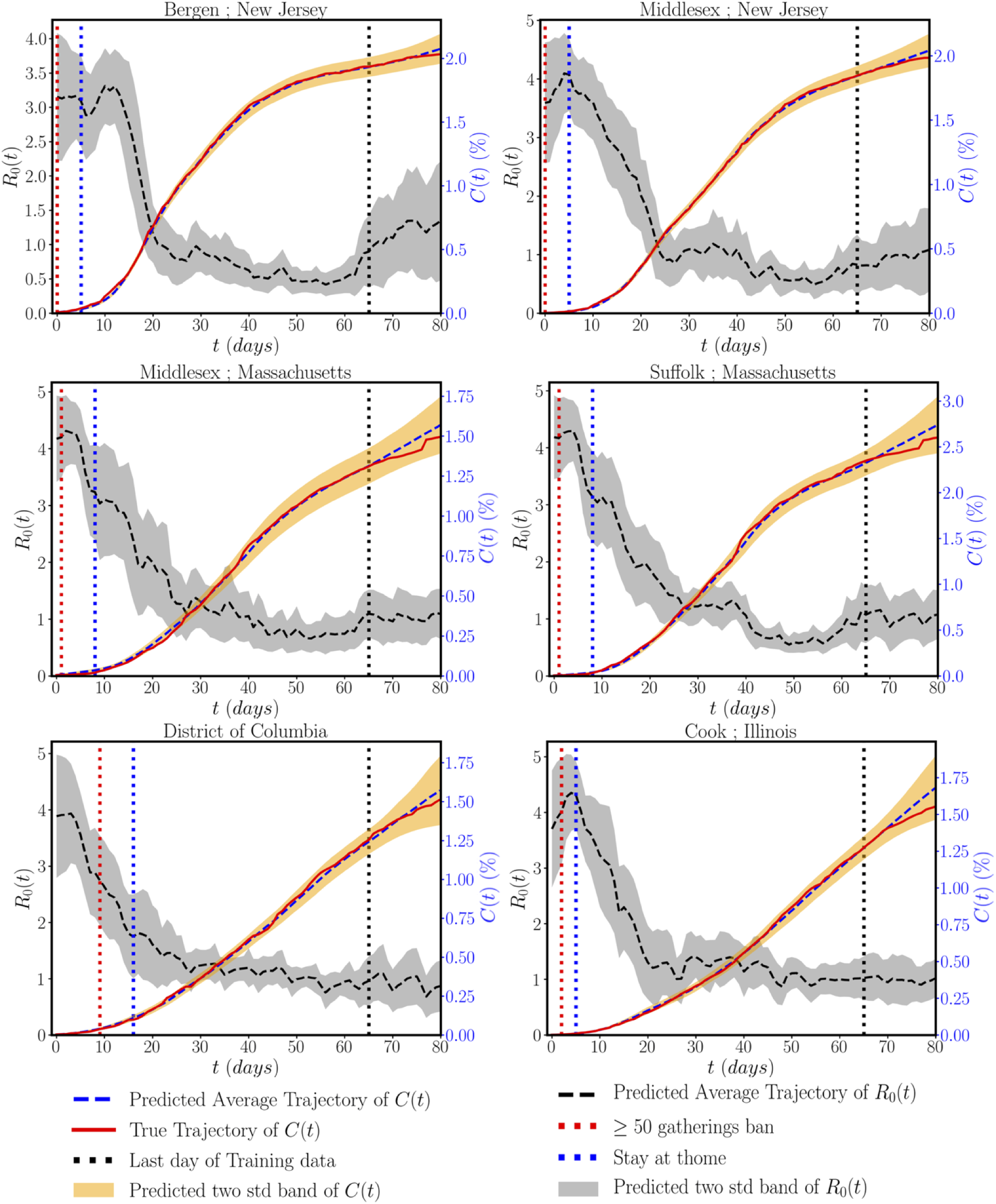
Evolution of the fraction of cumulative cases *C*(*t*) and the basic reproduction number *R*_0_(*t*) for representative counties across the US. The red and blue horizontal dashed lines indicate the dates corresponding to the large crowd gathering ban and stay at home order issued by each county. The black horizontal dashed line delineates the data used for training the model (days 0-65), and the predicted model forecasts (days 66-80).

**Figure 7:**
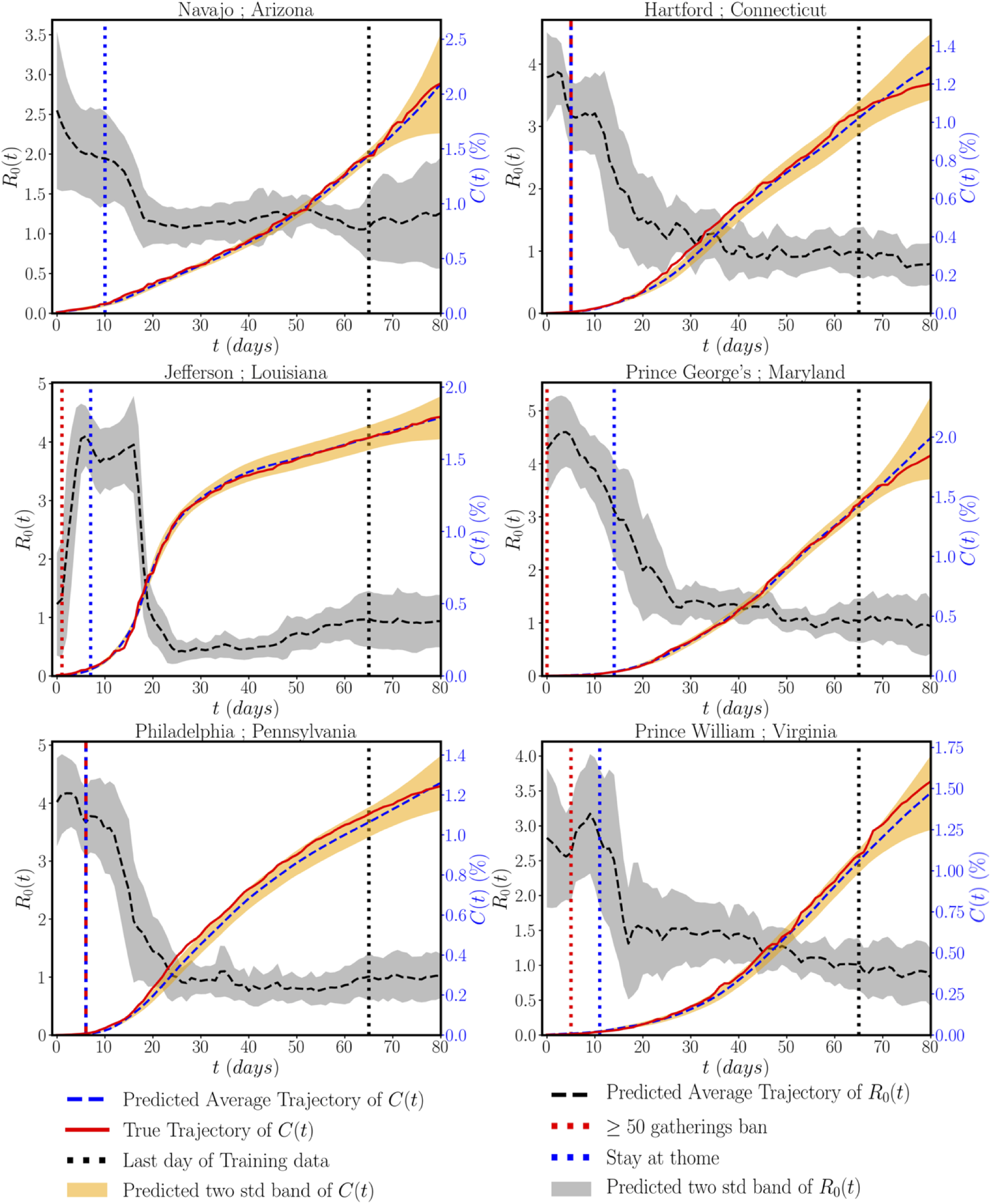
Evolution of the fraction of cumulative cases *C*(*t*) and the basic reproduction number *R*_0_(*t*) for representative counties across the US. The red and blue horizontal dashed lines indicate the dates corresponding to the large crowd gathering ban and stay at home order issued by each county. The black horizontal dashed line delineates the data used for training the model (days 0-65), and the predicted model forecasts (days 66-80).

### 3.3 Sensitivity Analysis

In order to assess the influence of different mobility and social behavior parameters on the evolution of the susceptible to infectious transition rate *β*, a sensitivity analysis using the Morris method [33] is performed on the trained LSTM neural network predicting 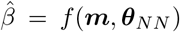. Note that the LSTM has *N*_*m*_ × *τ* inputs, such that 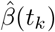 inference is carried out using ***m***(*t*_*k τ*_), …, ***m***(*t*_*k*−1_), where we remind the reader that 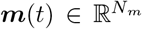 contains the mobility and social behavior parameters 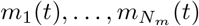. We use *s*_*i,j*_ to denote the modified mean of the Morris method sensitivity measure [34] of 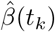 to *m*_*i*_(*t*_*k*−*j*_).

Figure 8 presents a heat-map of the normalized sensitivity measures *s*_*i,j*_. Strikingly, in view of the Google mobility parameters (*m*_1_, …*m*_6_), these results suggest that the sensitivity of *β* to parks attendance (parameter *m*_3_) — the only open place that is considered among the six available mobility parameters — is the lowest amongst all six. The second least influential parameter is the transit station’s attendance (parameter *m*_4_), reflective of a location where people spend on average the least amount of time among the places considered in the remaining five mobility parameters (except parameter *m*_3_). The numerical values obtained are also consistent with the COVID-19 incubation period estimated to be between 2 and 21 days by the United States’ CDC [35]. Indeed, the mobility parameters do not instantaneously affect *β* and a lag period should be taken into account in order to observe the effects of the mobility parameters on the spread of the disease, which corresponds to the incubation period and the testing delay. Regarding the Unacast social behavior parameters, all parameters display a significant sensitivity, with unnecessary visits displaying the highest value as shown in Figure 8.

**Figure 8:**
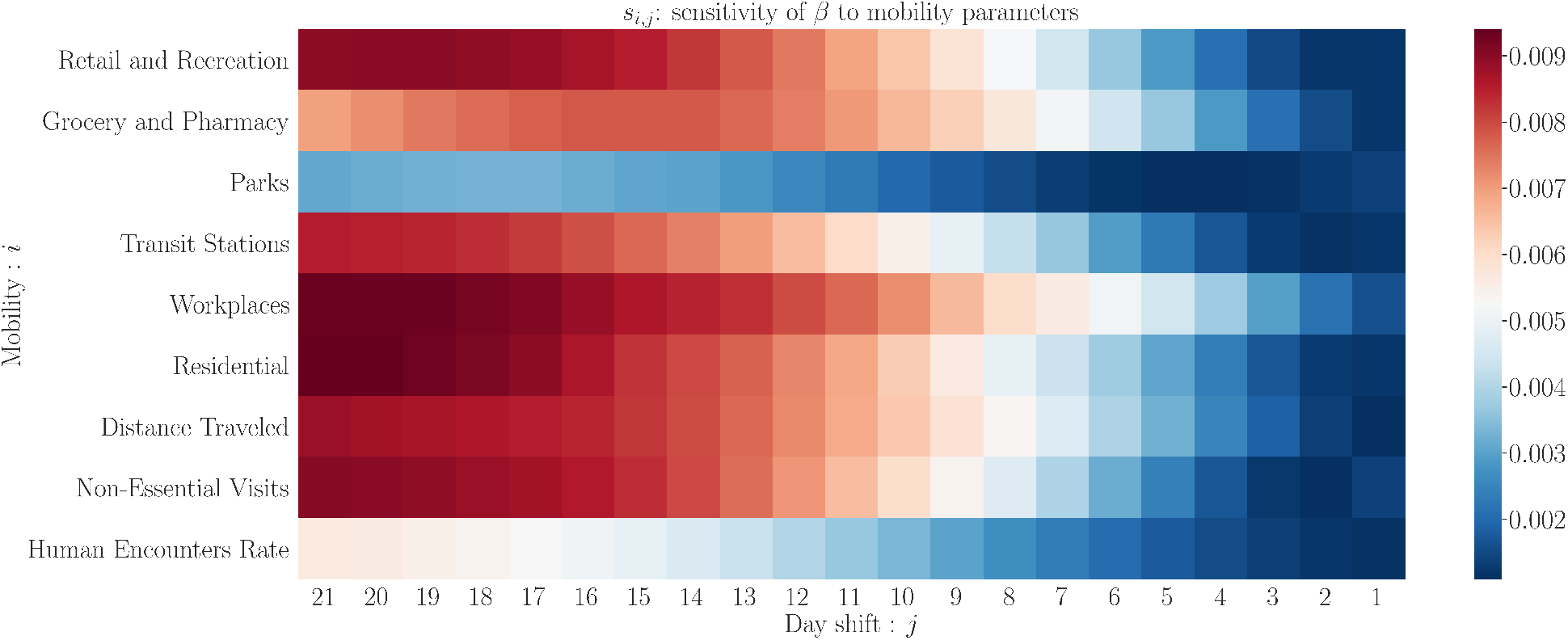
*s*_*i,j*_: Sensitivity of susceptible to infectious transition rate *β* to the mobility and social behavior parameters.

We also perform a local sensitivity analysis as follows. We increase each of the mobility and social behavior parameters by 10% for the last 21 days (each at a time), and we determine the corresponding prediction for the susceptible to infectious transition rate *β*. We then compare the obtained value for *β* for the last day to the actual one obtained with the unchanged data. Finally, we pick the parameter that corresponds to the highest change in *β*. This result, shown in Figure 9, allows us to locally identify the most sensitive parameter among the mobility and social behavior ones considered for each of the 203 counties studied. Interestingly, our results suggest that the spread of COVID-19 among a population of over 60 million people is most sensitive to the closure and reopening of workplaces, followed by changes in visits to residential places.

**Figure 9:**
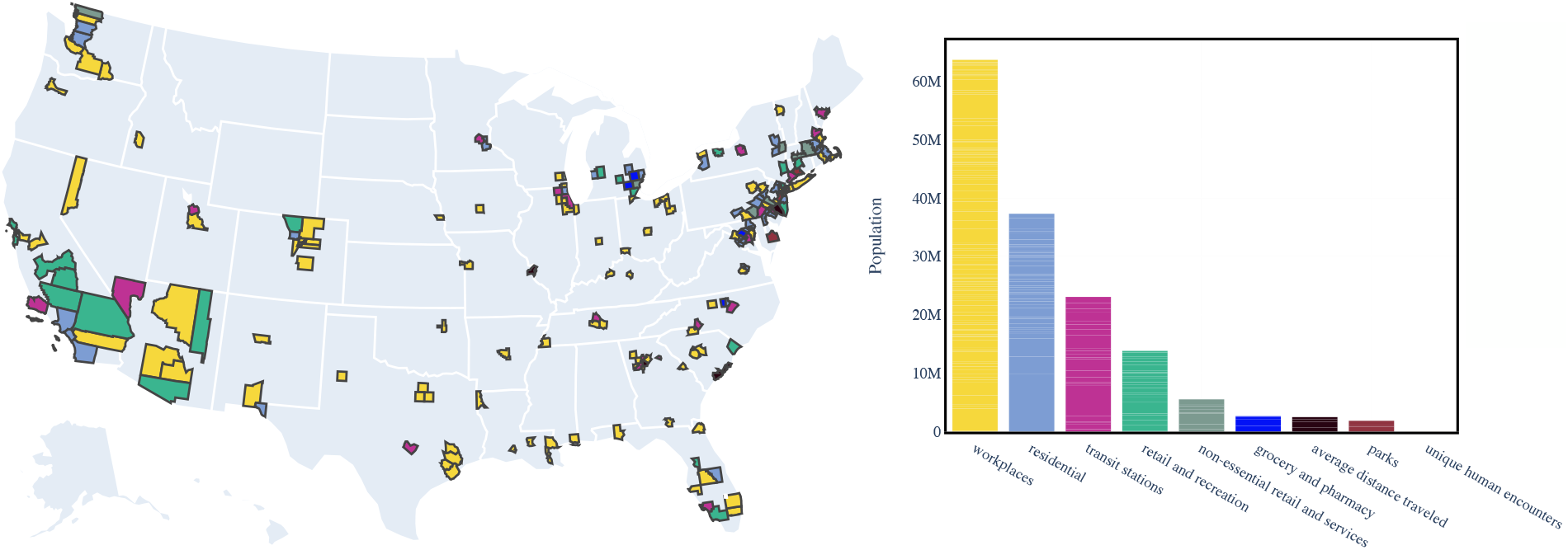
Mobility and social behavior parameters with the greatest effect on the reproduction number, by county (left panel) and by population (right panel).

### 3.4 Effects of Lock-down

In figures 5, 6 and 7 we indicated the day on which the *≥* 50 gatherings ban was issued along with the stay at home order day (except for Navajo, Arizona for which the *≥* 50 gatherings ban was issued at day *t* = − 4). The effect of these interventions on the evolution of the basic reproduction number *R*_0_(*t*) is significant, as the latter generally drops following at least one of these two policy decisions, such that *R*_0_(*t*) always reaches values around 1 while these decisions are still in place. We should note that banning *≥* 50 gatherings was sufficiently effective to decrease *R*_0_ in some locations, including most of New York counties; Massachusetts counties; and Prince George’s, Maryland, for instance. However, for other counties, the drop of *R*_0_ was significant only after issuing the stay at home order as observed for Suffolk, New York; Middlesex, New Jersey; Cook, Illinois; Jefferson, Louisiana; and Prince Williams, Virginia. These observations quantify the importance of limiting gatherings in managing COVID-19 spread and the necessity of issuing the stay at home orders in locations where gatherings bans alone are not enough to manage the spreading.

### 3.5 Effects of Re-opening

To quantify the effect of re-opening, the retail and recreation mobility parameter of New York county (New York) is artificially altered as shown in top left plot of Figure 10 (while the other mobility and social behavior parameters are kept the same). As shown in top right and bottom left plots of Figure 10, such policy has a significant effect on the evolution of *β*(*t*) and by consequence on the basic reproduction number *R*_0_(*t*) increased beyond 1 for the last 20 days of the simulation. As a result, the number of cumulative cases *C*(*t*) would significantly increase as shown in the bottom right plot of Figure 10 and the spread of the disease would be accelerated due to the increase of the retail and recreation mobility parameter. We note that the reproduction number is significantly affected only after 20 days of increasing the mobility parameter. This result is in line with the sensitivity analysis in Figure 8, which shows that the reproduction number is mostly affected by mobility between 17 and 21 days earlier. In practice, this delay poses a challenge to policy makers, which can only evaluate the effects of easing and imposing restrictions after two to three weeks.

**Figure 10:**
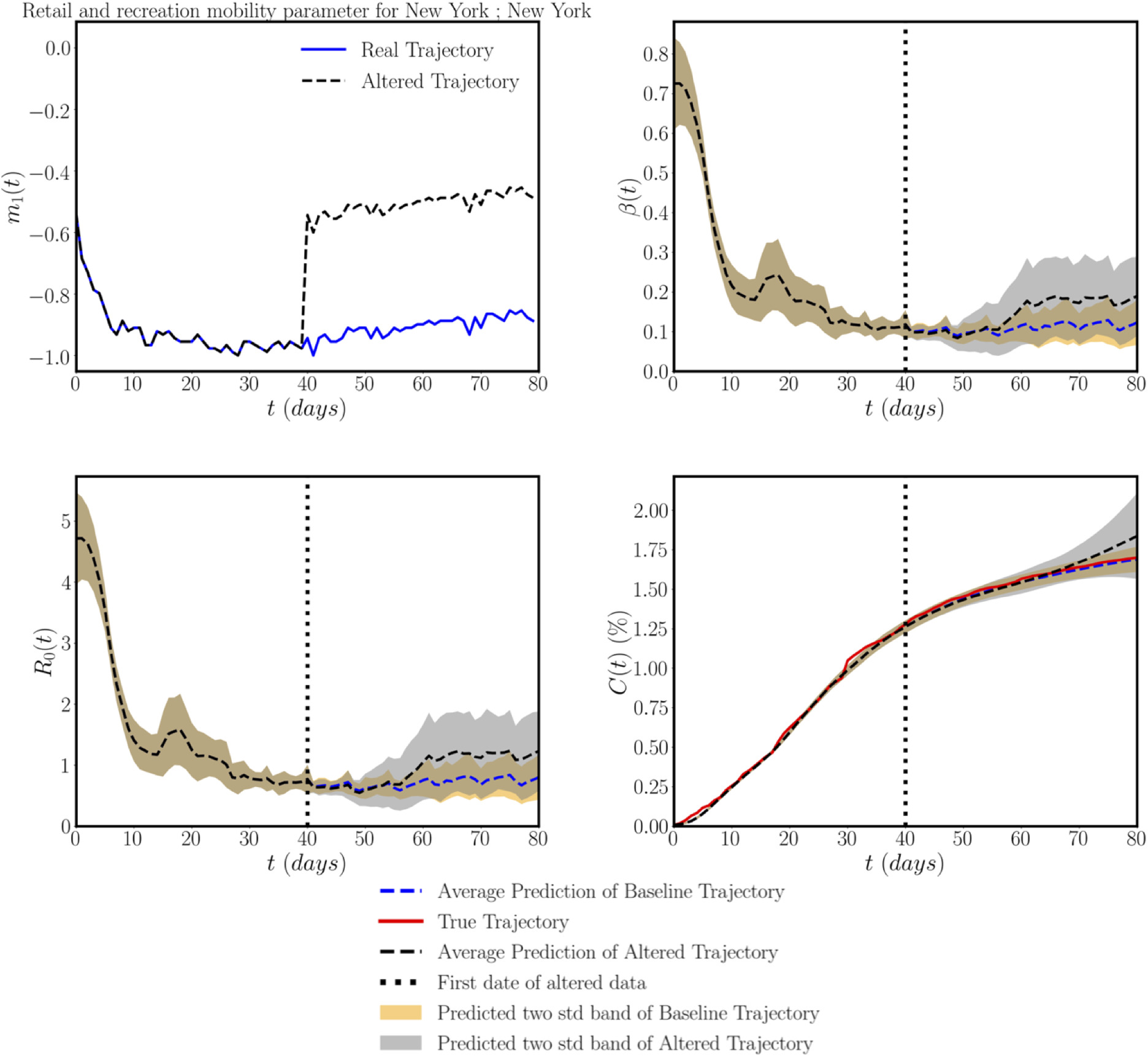
*Effects of re-opening*: Real and altered workplaces mobility parameter (top left), baseline and altered estimates of *β*(*t*) (top right), predicted basic reproduction number *R*(*t*) (bottom left), and cumulative cases percentage *C*(*t*) (bottom right).

## 4 Discussion

Amidst the COVID-19 outbreak, the modeling of the virus spread takes on life and death importance, as epidemic simulations shape national responses [36]. The reliability of such models relies heavily on their calibration on a set of incomplete and imperfect data points; a task that is often viewed as art and has been the source of scientific controversy [37]. In this work, we aspire to develop state-of-the-art machine learning algorithms to accurately determine the dynamic parameters characterizing the evolution of an epidemic, such as the time-dependent variation of the infectious rate and the basic reproduction number. Taking a systematic approach, the proposed model calibration framework will provide a quantitative understanding of the effects and efficiency of various policies designed to contain the spread of COVID-19 (and other infectious diseases) [38], and produce probabilistic forecasts for the evolution of a pandemic that can judiciously inform policy and decision making [39, 40].

Specific to this study, the proposed approach allows us to understand the most relevant mobility and social behavior parameters that affect the spread of COVID-19. Our results suggest that working from home policies have the significant effect in containing the reproduction number. This is shown in both global and local sensitivity analyses, where the mobility parameter that quantifies the amount of trips to workplaces is the most relevant among all mobility and social behavior parameters. Working in confined spaces has been shown to be a risk factor for the transmission of COVID-19 [41]. On the other hand, variations in park visits showed little influence in the dynamics of pandemic, which is in line with the reduced risk of transmission while outdoors [42]. A somehow surprising result is the mild influence of the human encounters rate parameter on the reproduction number. This could be explained by the use of personal protective equipment, such as face masks, which greatly reduces the risk of transmission and by a behavior change where human encounters moved from indoor to outdoor circumstances [43]. From these results, it appears that not all human encounters lead to an increased transmission, but only those that occur indoors and for a prolonged period of time, such as in workplaces and in residential visits.

Another relevant point that our analysis revealed is the delay between changes in mobility and social behavior and changes in the reproduction number. Variations that we see today in the reproduction number are mostly influenced by changes in behavior that occurred between 17 and 21 days ago. This time delay can be explained by the incubation period of the virus [35] and by the delay in obtaining and reporting positive test results. Without any model to relate mobility and social behavior to the pandemic dynamics, policy makers are posed with a real challenge of making decisions that will have a measurable outcome only after 2 to 3 weeks. This is where tools like ours can have an impact to guide and accelerate the decisions of opening or closing certain areas. We highlight the utility of our model by simulating an increase in retail and recreation mobility in New York county, for which we observe that the reproduction number starts increasing only after 20 days of the intervention. By using the proposed model, authorities could detect the threat of increased mobility early and act in time to prevent a new outbreak.

Our methodology presents some limitations that open new research opportunities. First, there are pronounced differences in the testing frequency both in space and time. These disparities will affect the number of detected cases and bias the data that we use to train our model. Including the number of tests performed as part of the input to our model could improve our predictions and assess the effect of limited testing capabilities. However, these kind of data are currently not available at the county level. Secondly, we have not included a potentially large fraction of cases that remain undetected or asymptomatic. This population can have an influence on the model behavior, especially when the case numbers are relatively large. Nonetheless, estimations of the undetected population based on antibody studies are currently available only at very few locations [24]. For this reason, we decided to not explicitly model this population, as we would be simply guessing its size. Finally, our model tends to perform relatively better in counties with more cases, as we can see in Figure 4. This is intrinsically linked to the loss function that we chose to train the model. In our case, this function tends to penalize more counties with more cases. We think this behavior is desirable because it makes our predictions more accurate in regions where the pandemic is rapidly evolving. Nonetheless, it would be straightforward to adjust the loss function to emphasize other critical factors in disease transmission if needed.

## Data Availability

The data that we used can be found as follows:
Google mobility data: https://www.google.com/covid19/mobility/
Accessed: June 8th 2020;
Unacast social behavior data:
https://www.unacast.com/data-for-good
Version from June 11th 2020.
The data obtained from our study will be made public once the paper is published.

## Acknowledgements

This work received support from the US Department of Energy under the Advanced Scientific Computing Research program (grant DE-SC0019116), the Defense Advanced Research Projects Agency under the Physics of Artificial Intelligence program (grant HR00111890034), and the Air Force Office of Scientific Research (grant FA9550-20-1-0060). M.P. and E.K. also acknowledge support from a Stanford Bio-X IIP Seed Grant, E.K. by the NIH Grant U01 HL119578, and K.L. by a DAAD Fellowship.

## Authors’ Contribution

M.A.B., P.P. and F.S.C. conceived the computational methods, M.A.B. implemented the methods, processed the data and performed the simulations, K.L, M.P. and E.K. proposed the epidemiology model and data, all authors wrote the manuscript.

## A Appendix 1: Loss Function and Multi-step Method

The forward pass of the LSTM neural network considered in this work for one single evaluation of 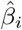 (predicted value of *β* at time instance *t*_*i*_) is given in Algorithm 1, where *σ* refers to the sigmoid function, ∘ to the element wise multiplication between two tensors of same shape and (*·, ·*) to the euclidean inner product between two vectors of same size. Based on the notation used in Algorithm 1, the LSTM parameters ***θ***_*NN*_ can be expressed as follows:

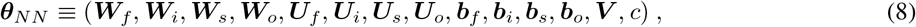

where ***W***_*f*_, ***U***_*f*_ ***b***_*f*_ refer to the forget gate’s parameters, ***W***_*i*_, ***U***_*i*_ ***b***_*i*_ to the input/update gate’s parameters, ***W***_*s*_, ***U***_*s*_ ***b***_*s*_ to the cell state gate’s parameters, ***W***_*o*_, ***U***_*o*_ ***b***_*o*_ to the output gate’s parameters and ***V***, *c* to the output layer parameters. *d*_*h*_ refers to the hidden state dimension and will be taken equal to 120. These parameters are initialized following the Xavier method [44].

Figure 12 shows the convergence of the loss function of Eq (7) with the number of iterations. Other loss functions have been investigated, including:

**Figure 11:**
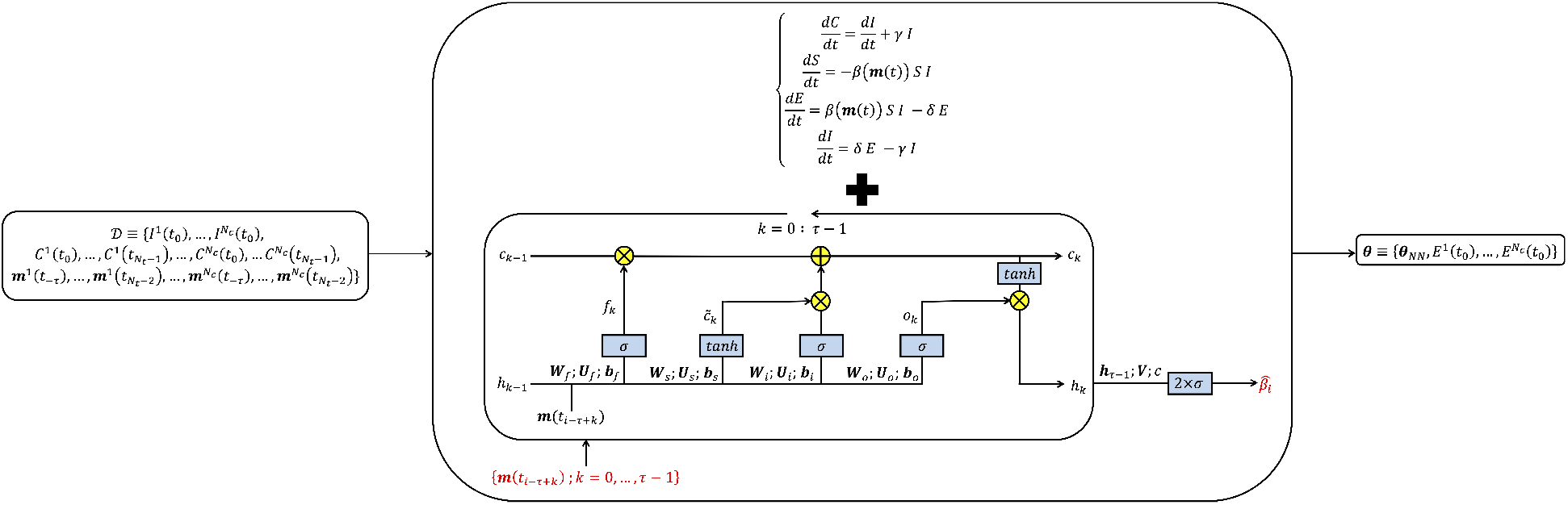
*Methodology summary:* Using the dataset 𝒟 and the mobility and social behavior-based SEIR model, the multi-step LSTM parameters along with the initial fractions of the exposed population for the different counties are estimated by minimizing the loss function (7)

**Figure 12:**
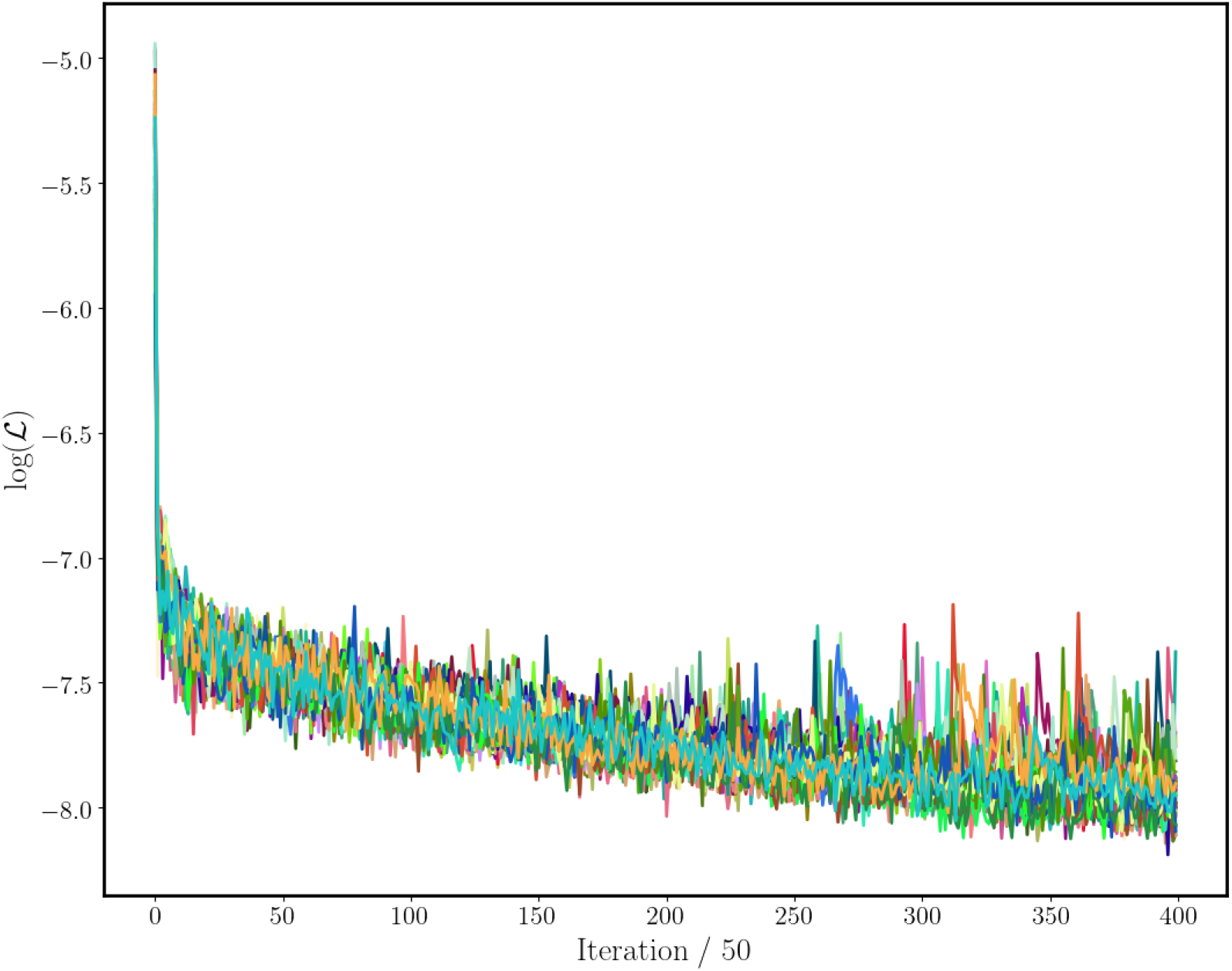
*Convergence of training loss* (Logarithm of ℒ in Eq (7))

1. a loss based on the residual of the differential equation of *I*(*t*) in the SEIR model satisfied by *Î*(*t*),
2. a loss based on the *L*_2_ difference between the data of *C*(*t*) and the inferred number of cumulative cases: 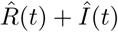
3. a loss based on the *L*_2_ difference between the increase of number of cumulative cases *C*(*t*_*i*_) − *C*(*t*_*i*−1_) and the inferred number of such increase 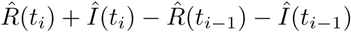

The loss defined in (7) performs better than the loss mentioned in point 1 because the latter is more prone to local minima if it is not initialized sufficiently close to the exact solution and due to the noisy data and numerical errors, the proposed loss (7) gave considerably better results. The losses in points 2 and 3 require one additional time integration to estimate the variable *R*(*t*). Hence, such step introduces additional numerical errors, which explains the better performance observed with the loss defined in (7).

### Algorithm 1: LSTM forward pass

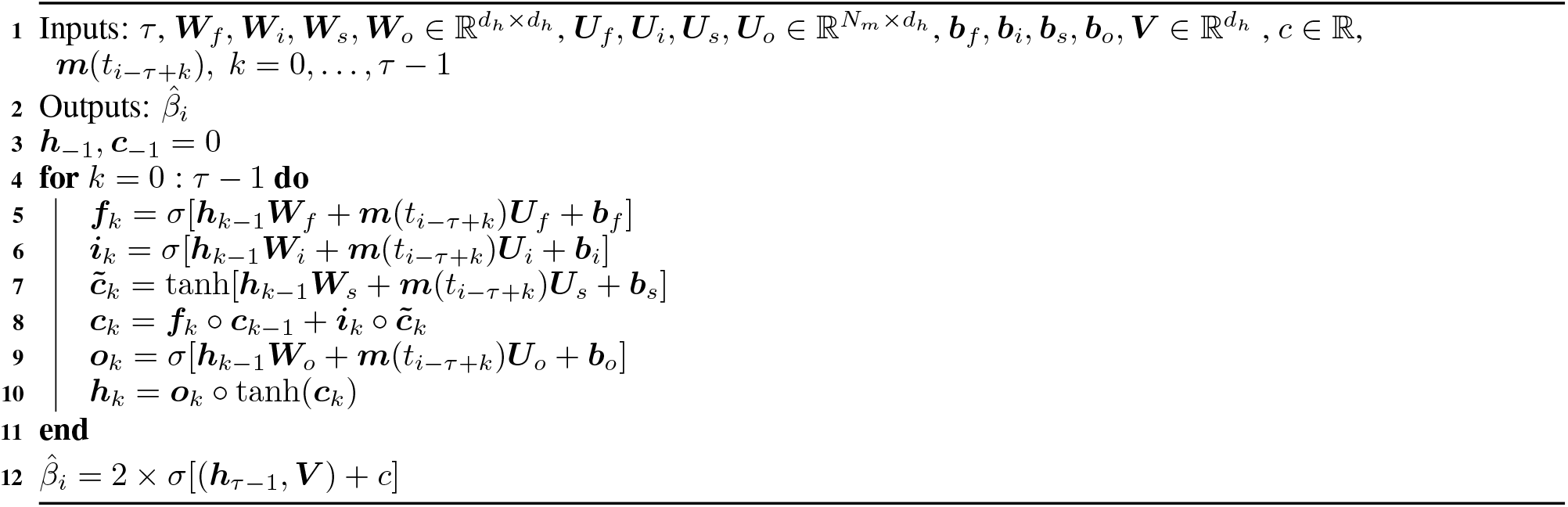

Various multistep methods were investigated including implicit Adams-Moulton (AM) and explicit Adams-Bashford (AB) schemes with different orders. The implicit AM schemes perform better than the explicit AB ones thanks to their numerical stability. Higher order schemes resulted in stiffer numerical systems which sometimes led to numerical instabilities during model training. Therefore, the first order AM scheme (trapezoidal scheme) was adopted in this work.

## B Appendix 2: Comparison of predictive accuracy against a simple SEIR model with a constant reproduction number *β*

The same model described in by equation (1) is adopted to utilize the cumulative data. In this case, the value of *γ* and *δ* are assumed as given and equal to 1*/*6.5 [27, 28] and 1*/*2.5 [27, 11] as previous. We assume that the parameter *β* varies from one county to another but is fixed over time and independently distributed with respect to some prior distribution. In this case the prior distribution is a truncated normal distribution with mean equal to 0.5 and a scale of 0.5. Furthermore, for every county, we assume that each entry of log *C*, i.e., the natural logarithm of cumulative case fraction, follows i.i.d normal distributions centered at log(*Ĉ*), with their standard deviation following a log normal distribution with mean 0 and scale 10. Given such prior, the model is sampled using the NUTS sampler from Numpyro package [45] for 4, 500 iterations, with a target acceptance probability of 0.85. We then use the mean of the last 3, 000 samplings as an estimator for the parameter *β*. The relative error averaged over the extrapolation period obtained with such model is shown in the right plot of Figure 13 for the different counties. We also tried several different priors for *β*, but all of them gave similar results. In general, the simple SEIR model tends to produce cumulative cases fraction variations that are convex within the period of time considered, while the actual trajectories generally change concavity, due to the enforced behavioral intervention policies, such as lock-down and gatherings ban.

## Footnotes

1 Code available at https://github.com/PredictiveIntelligenceLab/DeepCOVID19.

## Notes

### Competing Interest Statement

The authors have declared no competing interest.

### Author Declarations

no IRB needed

